# Data Anonymization for Open Science: A Case Study

**DOI:** 10.1101/2024.12.16.24319068

**Authors:** Paul Francis, Gregor Jurak, Bojan Leskošek, Karen Otte, Fabian Prasser

## Abstract

One of many challenges to open science is anonymization of personal data so that it may be shared. This paper presents a case study of the anonymization of a dataset containing cardio-respiratory fitness and commuting patterns for Slovenian school children. It evaluates three different anonymization tools, ARX, SDV, and SynDiffix. The fitness study was selected because its small size (N=713) and generally low statistical significance make it particularly challenging for data anonymization. Unlike most prior anonymization tool evaluations, this paper examines whether the scientific conclusions of the original study would have been supported by the anonymized datasets. It also considers the burden imposed on researchers using the tools both for data generation and data analysis.

## Introduction

A core function of open science is data sharing. In an ideal setting, scientists should be able to easily upload data to a repository which satisfies the FAIR principles, i.e. makes the data findable, accessible, interoperable, and resuable [1]. Data sharing is challenging even when the data is not personal data [2], i.e. does not contain information about individuals. Sharing of personal data is even more challenging [3] due to stricter and more diverse legal frameworks. In a data sharing pipeline where personal data is involved, there is often an extra step of data anonymization that historically has required individual attention to each data release, for instance in order to find a good balance between privacy and utility [4]. At the same time, there are constant advances in data anonymization technology. In recent years for instance, a new generation of synthetic data tools have become available[5], both as open source and commercial products.

This study assesses the applicability of a small but representative set of data anonymization tools to an open science personal data sharing scenario. For this, we selected a *base study*, which we tried to replicate using anonymized data. The base study is a research study authored by GJ and BL [6] with data controlled by them. The study, titled *“Associations of mode and distance of commuting to school with cardiorespiratory fitness in Slovenian school children: a nationwide cross-sectional study”*, is challenging for data anonymization tools due to its small sample size (713 records), the amount and depth of the statistical analyses, and overall few significant results and small effect sizes. These conditions are challenging because the distortion necessarily introduced by anonymization must be small so as not to overwhelm the significance of the data.

The evaluation in this study focuses on three questions:

1. Would the scientific conclusions of the study have changed if anonymized data were used instead of the original data?
2. How difficult would it have been to use the anonymized data for the scientific study?
3. How difficult would it have been for non-experts to generate the anonymized data?

The three anonymization tools evaluated are ARX [7] (developed by Prasser), SynDiffix [8] (developed by Francis), and SDV [9]. All tools are open source software and implement different methods to create anonymized datasets. More information about these tools, as well as the procedure used to select them, can be found in the Methods section of this paper.

## Results

The base study of Jurak et al. investigated whether active commuting has the potential to improve children’s health[6]. The cardiorespiratory fitness (CRF) of 713 Slovenian school children aged 12 to 15 years, was determined with a 20-m shuttle run test to estimate their maximal oxygen uptake (VO_2_max). Moreover, information was collected on their distance from home to school and whether the commute was done by walking, wheels (e.g., bicycle or skateboard), public transport or car in both directions. The study found that commuting distance minimally affected CRF, except in the Car group, where children living closer to school had significantly lower CRF than those farther away. The study recommends targeting car-driven children within walking or cycling distance of school with interventions promoting active transport. The original paper presented its results in three tables and one figure which we refer to as base-tables and base-figure. This study replicates these base-tables and base-figure, but in such a way that the the original data and the data for the three anonymized datasets are combined for easy comparison.

The following sections discuss the similarities and differences between the anonymized and original data, and comment on the extent to which the conclusions of the original paper still hold given the anonymized data.

### Commute modes and distances (Base-tables 1 and 2 from the original paper)

The original and anonymized data for base-tables 1 and 2 explored the types of commuting in school children and the commute distances traveled. These are given in this paper’s Tables 1 and 2 respectively.

**Table 1.**
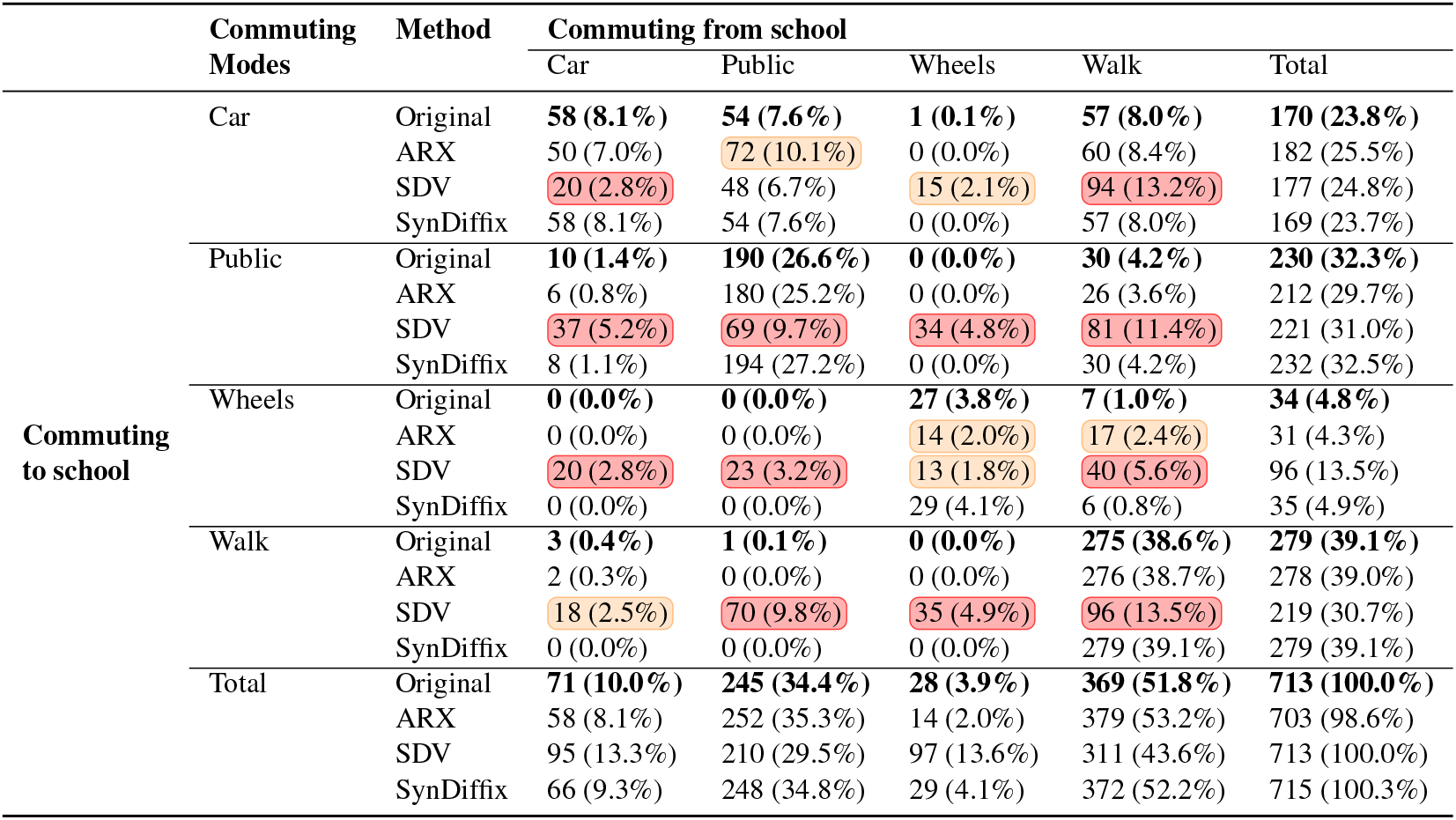
Base-table 1 from the paper showing the counts and percentages for the original data and the three anonymization methods. Each group of four presents the data in order of Original, ARX, SDV, and SynDiffix. Counts and their corresponding percentages are shaded 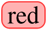 when the absolute error is greater than 20 or the relative error is greater than 30%. They are shaded 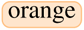 when the absolute error is greater than 10 or the relative error is greater than 15%.

|  | Commuting Modes | Method | Commuting from school |  |  |  |  |
| --- | --- | --- | --- | --- | --- | --- | --- |
|  |  |  | Car | Public | Wheels | Walk | Total |
| Commuting to school | Car | Original | **58 (8.1%)** | **54 (7.6%)** | **1 (0.1%)** | **57 (8.0%)** | **170 (23.8%)** |
|  |  | ARX | 50 (7.0%) | 72 (10.1%) | 0 (0.0%) | 60 (8.4%) | 182 (25.5%) |
|  |  | SDV | 20 (2.8%) | 48 (6.7%) | 15 (2.1%) | 94 (13.2%) | 177 (24.8%) |
|  |  | SynDiffix | 58 (8.1%) | 54 (7.6%) | 0 (0.0%) | 57 (8.0%) | 169 (23.7%) |
|  | Public | Original | **10 (1.4%)** | **190 (26.6%)** | **0 (0.0%)** | **30 (4.2%)** | **230 (32.3%)** |
|  |  | ARX | 6 (0.8%) | 180 (25.2%) | 0 (0.0%) | 26 (3.6%) | 212 (29.7%) |
|  |  | SDV | 37 (5.2%) | 69 (9.7%) | 34 (4.8%) | 81 (11.4%) | 221 (31.0%) |
|  |  | SynDiffix | 8 (1.1%) | 194 (27.2%) | 0 (0.0%) | 30 (4.2%) | 232 (32.5%) |
|  | Wheels | Original | **0 (0.0%)** | **0 (0.0%)** | **27 (3.8%)** | **7 (1.0%)** | **34 (4.8%)** |
|  |  | ARX | 0 (0.0%) | 0 (0.0%) | 14 (2.0%) | 17 (2.4%) | 31 (4.3%) |
|  |  | SDV | 20 (2.8%) | 23 (3.2%) | 13 (1.8%) | 40 (5.6%) | 96 (13.5%) |
|  |  | SynDiffix | 0 (0.0%) | 0 (0.0%) | 29 (4.1%) | 6 (0.8%) | 35 (4.9%) |
|  | Walk | Original | **3 (0.4%)** | **1 (0.1%)** | **0 (0.0%)** | **275 (38.6%)** | **279 (39.1%)** |
|  |  | ARX | 2 (0.3%) | 0 (0.0%) | 0 (0.0%) | 276 (38.7%) | 278 (39.0%) |
|  |  | SDV | 18 (2.5%) | 70 (9.8%) | 35 (4.9%) | 96 (13.5%) | 219 (30.7%) |
|  |  | SynDiffix | 0 (0.0%) | 0 (0.0%) | 0 (0.0%) | 279 (39.1%) | 279 (39.1%) |
|  | Total | Original | **71 (10.0%)** | **245 (34.4%)** | **28 (3.9%)** | **369 (51.8%)** | **713 (100.0%)** |
|  |  | ARX | 58 (8.1%) | 252 (35.3%) | 14 (2.0%) | 379 (53.2%) | 703 (98.6%) |
|  |  | SDV | 95 (13.3%) | 210 (29.5%) | 97 (13.6%) | 311 (43.6%) | 713 (100.0%) |
|  |  | SynDiffix | 66 (9.3%) | 248 (34.8%) | 29 (4.1%) | 372 (52.2%) | 715 (100.3%) |

**Table 2.** Base-table 2 from the original paper showing the counts and distances in meters (median and IQR) for the original data and the three anonymization methods. Each group of four presents the data in order of Original, ARX, SDV, and SynDiffix. The shading for counts (N) are as described for Table 1. Distance and IRQ are shaded 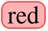 when the relative error is greater than 30%, and shaded 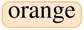 when the relative error is greater than 15%. Note that the original distances median and IQR don’t perfectly match those of the original Table 2 because of differences in the way median and IQR were calculated (Python versus R).

| Commuting group | Method | From home to school |  | From school to home |  |
| --- | --- | --- | --- | --- | --- |
|  |  | N (%) | Distance (IQR) | N (%) | Distance (IQR) |
| Car | Original | **170 (24%)** | **3133 (3945)** | **71 (10%)** | **3615 (3896)** |
|  | ARX | 182 (26%) | 3758 (3915) | 58 (8%) | 3910 (3800) |
|  | SDV | 177 (25%) | 7602 (8467) | 95 (13%) | 3934 (7362) |
|  | SynDiffix | 169 (24%) | 3754 (4215) | 70 (10%) | 2584 (3560) |
| Public | Original | **230 (32%)** | **4782 (4296)** | **245 (34%)** | **4996 (4033)** |
|  | ARX | 212 (30%) | 4973 (4193) | 252 (35%) | 5140 (3686) |
|  | SDV | 221 (31%) | 5690 (8320) | 210 (29%) | 2249 (5174) |
|  | SynDiffix | 232 (33%) | 4712 (4228) | 245 (34%) | 5011 (4097) |
| Wheels | Original | **34 (5%)** | **1366 (2211)** | **28 (4%)** | **1444 (2369)** |
|  | ARX | 31 (4%) | 1356 (1378) | 14 (2%) | 2235 (3245) |
|  | SDV | 96 (13%) | 6671 (8472) | 97 (14%) | 2741 (5282) |
|  | SynDiffix | 36 (5%) | 1094 (2251) | 30 (4%) | 1337 (1376) |
| Walk | Original | **279 (39%)** | **799 (789)** | **369 (52%)** | **973 (1043)** |
|  | ARX | 278 (39%) | 805 (795) | 379 (53%) | 954 (1062) |
|  | SDV | 219 (31%) | 5498 (8697) | 311 (44%) | 2374 (6068) |
|  | SynDiffix | 279 (39%) | 787 (737) | 368 (52%) | 960 (1037) |
| Total | Original | **713 (100%)** |  | **713 (100%)** |  |
|  | ARX | 703 (99%) |  | 703 (99%) |  |
|  | SDV | 713 (100%) |  | 713 (100%) |  |
|  | SynDiffix | 716 (100%) |  | 713 (100%) |  |

Among the three anonymization methods, SynDiffix most closely reproduced original counts and percentages for cross-tabulation of commuting modes (Table 1), with only one deviating record in the cross table. It is closely followed by ARX which showed up to 18 deviations, while SDV provides large differences of up to 179 different commute mode counts in the cross table. Nevertheless, the basic description of these results in original paper, i.e. that active commuting is more frequent in the direction from school to home than in other direction, still holds, but this may be a coincidence in case of SDV as it substantially over-or underestimates the frequencies for both modes of active commuting (walking and wheels). Note also that the anonymized table of ARX contains 1.4% fewer rows (703 versus 713) due to suppression of rare records.

Similar results were found for commuting distance (Table 2). Here also SDV performs badly, hugely over-or underestimating the distance in most of the table cells, both in central tendency (median) and variability (IQR) of the data. SynDiffix and ARX perform much better, but still with large errors in some of the table cells, especially in cells with low frequencies, e.g. SynDiffix substantially underestimates the median distance of car commuting from school to home (3615→ 2584) and ARX overestimates the distance of wheels commuting in same direction (1444 →2235). The verbal description of this results from original paper, i.e. that active commuting groups (Walk, Wheels) typically live close to school, still holds in case of SynDiffix and ARX, but not in case of SDV.

The absolute errors of the cross-table counts from Table 1 and the distance values in Tables from 2 are visualized in the boxplots of Figure 1.

**Figure 1.**
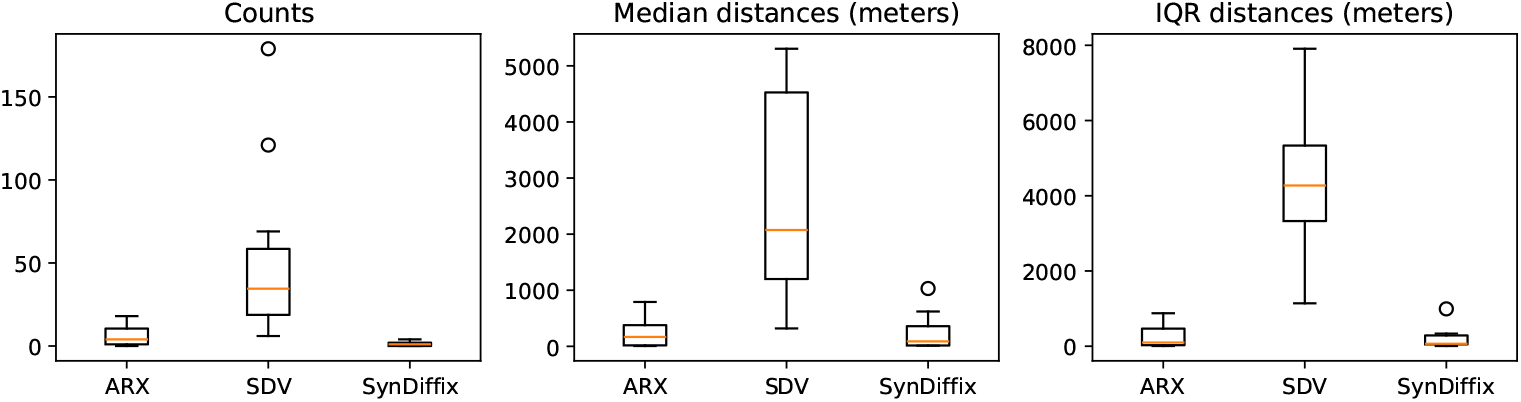
Absolute error of the three anonymization methods for the counts and distances in Tables 1 and 2. Each box plot data point is taken from a single cell in Tables 1 and 2.

### Statistical significance of regression coefficients (Base-table 3 from the original paper)

The original and anonymized data for base-table 3 are given is this paper’s Tables 3 and 4 (the data is spread over two tables for formatting purposes). The primary purpose of base-table 3 is to indicate statistical significance. As such, those entries where the original data is significant and the anonymized data is not, or vice versa, are highlighted in red. In total, there are 3 such mismatches for ARX, 8 for SDV, and 5 for SynDiffix.

**Table 3.** Part 1 (of 2) of the original paper’s Table 3 showing the parameters (regression coefficients) of the linear model for prediction of VO2max by group and distance. 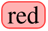 shading indcates that the anonymized entry is non-significant where the original data is significant or vice versa. Each group of four presents the data in order of Original (bold), ARX, SDV, and SynDiffix.

| Variables | Adjusted model |  |  |  |
| --- | --- | --- | --- | --- |
|  | From home to school |  | From school to home |  |
|  | Coefficient | 95% CI | Coefficient | 95% CI |
| **Constant** | **36.42\*\*\*** | **(28.17, 44.67)** | **36.63\*\*\*** | **(29.11, 44.15)** |
|  | 33.19*** | (25.82, 40.56) | 35.36*** | (28.65, 42.07) |
|  | 56.08*** | (45.07, 67.09) | 49.18*** | (40.44, 57.91) |
|  | 27.94*** | (20.74, 35.14) | 32.89*** | (26.25, 39.54) |
| **Commuting group** |  |  |  |  |
| Car | **-6.49** | **(-15.92, 2.94)** | **-15.13\*** | **(-26.88, -3.39)** |
|  | -7.28 | (-15.49, 0.92) | -17.72** | (-29.65, -5.8) |
|  | -9.17 | (-21.3, 2.95) | 2.4 | (-6.44, 11.24) |
|  | -2.18 | (-9.25, 4.88) | -12.67* | (-22.82, -2.52) |
| Public | **-0.08** | **(-9.06, 8.9)** | **-3.19** | **(-11.27, 4.88)** |
|  | 3.21 | (-4.59, 11.01) | -4.08 | (-10.99, 2.84) |
|  | -6.17 | (-16.67, 4.32) | -2.57 | (-9.06, 3.92) |
|  | -0.15 | (-6.37, 6.07) | 8.71** | (2.72, 14.71) |
| Wheels | **3.0** | **(-16.24, 22.25)** | **15.66** | **(-4.09, 35.41)** |
|  | 3.88 | (-11.83, 19.58) | 17.16 | (-4.9, 39.22) |
|  | -8.69 | (-23.52, 6.14) | 1.48 | (-7.47, 10.44) |
|  | 3.23 | (-10.68, 17.13) | 10.7 | (-3.6, 24.99) |
| Walk (ref) |  |  |  |  |
| **Interaction Commuting group x Distance** |  |  |  |  |
| Car x Distance | **0.58** | **(-0.04, 1.2)** | **1.25\*\*** | **(0.34, 2.17)** |
|  | 0.79** | (0.24, 1.34) | 1.38** | (0.44, 2.33) |
|  | 0.35 | (-0.35, 1.06) | -0.28 | (-0.94, 0.37) |
|  | 0.42 | (-0.08, 0.91) | 2.06*** | (1.19, 2.93) |
| Public x Distance | **0.06** | **(-0.49, 0.61)** | **0.33** | **(-0.21, 0.88)** |
|  | -0.04 | (-0.52, 0.45) | 0.37 | (-0.1, 0.84) |
|  | 0.04 | (-0.48, 0.56) | 0.38 | (-0.06, 0.82) |
|  | 0.26 | (-0.13, 0.65) | 0.03 | (-0.39, 0.45) |
| Wheels x Distance | **-0.09** | **(-1.79, 1.62)** | **-1.15** | **(-2.89, 0.6)** |
|  | 0.08 | (-1.32, 1.48) | -1.41 | (-3.35, 0.53) |
|  | 0.09 | (-0.88, 1.07) | -0.04 | (-0.75, 0.66) |
|  | 0.19 | (-1.07, 1.44) | -0.13 | (-1.36, 1.11) |
| Walk x Distance | **-0.02** | **(-0.62, 0.58)** | **0.03** | **(-0.42, 0.48)** |
|  | 0.17 | (-0.33, 0.68) | -0.04 | (-0.42, 0.34) |
|  | -0.63 | (-1.28, 0.02) | -0.08 | (-0.44, 0.28) |
|  | 0.17 | (-0.24, 0.58) | 0.66*** | (0.3, 1.02) |

\* p ≤ 0.05, \*\* p ≤ 0.01, \*\*\* p ≤ 0.001

**Table 4.**
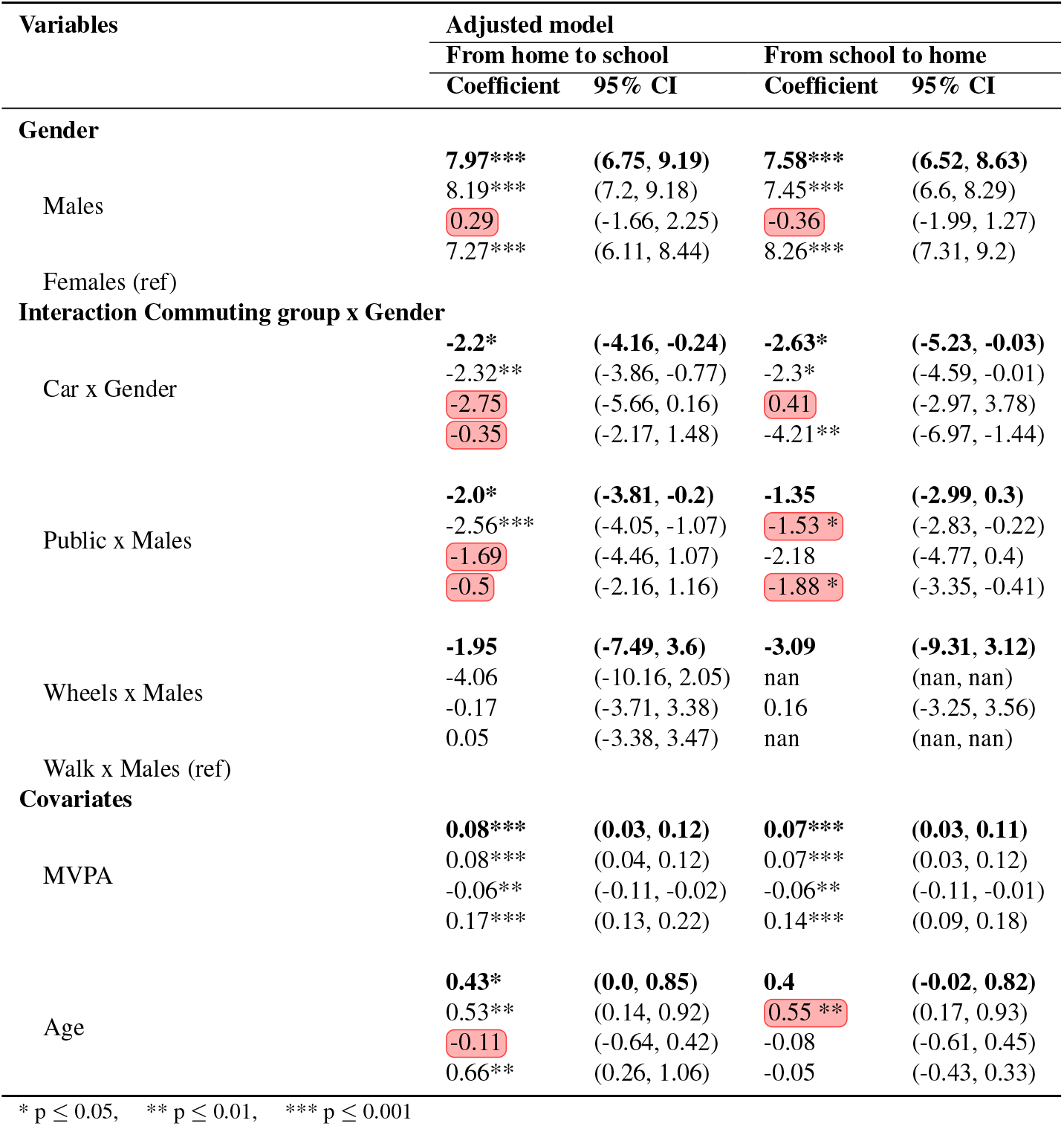
Part 2 (of 2) of the original paper’s Table 3 showing the parameters (regression coefficients) of the linear model for prediction of VO2max by group and distance. 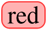 shading indcates that the anonymized entry is non-significant where the original data is significant or vice versa. Each group of four presents the data in order of Original (bold), ARX, SDV, and SynDiffix.

Several differences were found in statistics of linear prediction models based on original data and the data produced by all three anonymization methods (Tables 3 and 4). In the original models predicting VO2max, the constant (intercept), the predictors Gender and MVPA, and the derived Car × Gender interaction term were statistically significant in both directions of commuting. This also holds for all parameters in the SynDiffix and ARX models, except for the Car x Gender parameter in the SynDiffix model, which is not significant in one of the directions of commuting. The Gender parameter in the SDV model was found to be non-significant, despite being highly significant (p≤.001) in the three other models.

Note that only the SDV method provides estimate for Wheels x Males interaction parameter in the school to home direction models. ARX and SynDiffix suppressed this data as part of anonymization because there were so few datapoints. The normalized error for coefficients in Tables 3 and 4 is illustrated in Figure 2.

**Figure 2.**
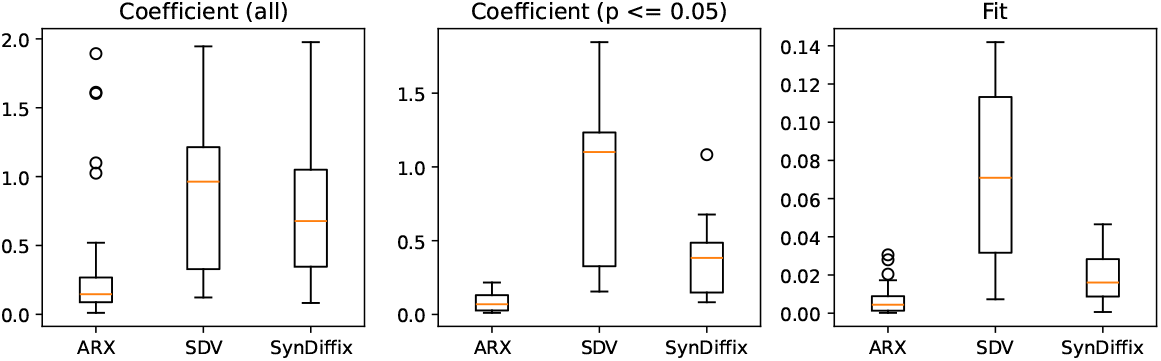
Normalized error for coefficients in Tables 3 and 4, and Fit (V0_2_max predicted at mode for median distance) for Figure 3. Normalized error *E* is computed as 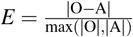,where *O* is the original value and *A* is the anonymized value. The middle table displays normalized error only for the datapoints in Tables 3 and 4 where the original coefficient is significant.

### Stratification of children’s cardiorespiratory fitness (Base-figure 1 from the original paper)

The plot for base-figure 1 is given in this paper’s Figure 3, which replicates the plot from the original paper as well as gives the corresponding plots for the three anonymized datasets. In male children, point predictions and prediction intervals in the original paper are quite closely matched with the ones from ARX and SynDiffix, but not with SDV. In female children, original statistics are most closely matched by ARX, followed SynDiffix and then by SDV. SDV gives similar results for male and female children, although they are clearly separated in the original data. SDV performs worst also in estimating prediction interval widths, which are very similar for both sexes, although being quite different in the original plot. ARX and SynDiffix produce interval widths that are more similar to original, but still differ in some cases, e.g. being too narrow in case of walk commuting in ARX and too narrow in females’ wheels commuting from school to home in both ARX and SynDiffix.

**Figure 3.**
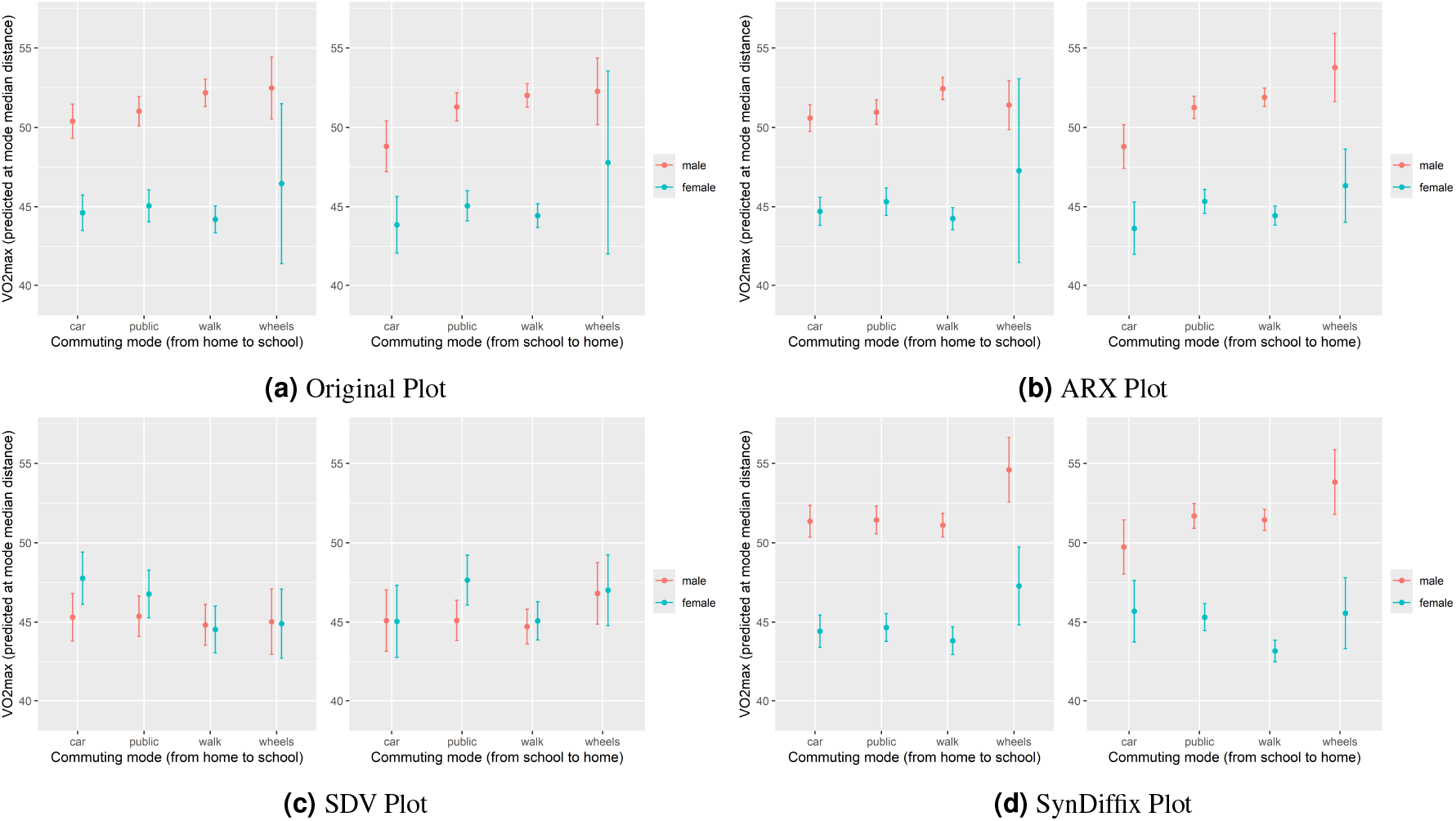
Comparison of the VO2max data plots corresponding to the base-figure from the original study.

### Comparison of derived scientific insights

Table 5 summarizes the ability of each anonymization method to produce the same analytic conclusions as those of the base paper. It presents the set of statements made in the base paper, and evaluates whether the statement would be supported 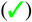, negated 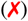, or neither supported nor negated 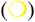.

**Table 5.** This table summarizes the ability of each anonymization method to result in the same analytic conclusion as that of the original data. 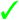 means that the correct conclusion is reached, 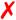 means that an incorrect conclusion is reached, and 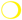 is inconclusive.

|  |  |  |  |
| --- | --- | --- | --- |
| **Main policy conclusion** | ARX | SDV | SynDiffix |
| Children driven by car who live within wheels or walk distance from school should be targeted by interventions promoting active transport | ✓ | ✗ | ✓ |
| **Descriptive analytics results** | ARX | SDV | SynDiffix |
| Overall, 43% of the participants reported active commuting modes to and from school and an additional 13% only in one direction | ✓ | ✗ | ✓ |
| Participants more often used active commuting from school (56%) than to school (44%) | ✓ | ✓ | ✓ |
| Males and females were choosing active commuting equally often | ✓ | ✓ | ✓ |
| Females preferred to walk and males preferred to use wheels transport | ✓ | ✗ | ✓ |
| The Walk group had the lowest and Public group had the highest median distance from home to school | ✓ | ✗ | ✓ |
| The active commuting groups (Walk Wheels) typically live close to school | ✓ | ✗ | ✓ |
| **CRF-related analytic results** | ARX | SDV | SynDiffix |
| When commuting from school to home, both the main effect of commuting group and its interaction with distance were significant | ✓ | ✗ | ✓ |
| The Car group had a significantly lower predicted value of CRF | ✓ | ✗ | ✓ |
| The Car group was the only one where distance of travel was related to CRF having significant difference to the reference (Walk) group; the participants with larger car travel distance had higher predicted VO2max | ✓ | ✗ | ○ |
| Overall interaction of commuting group with gender was not significant | ○ | ✓ | ○ |
| The largest sample difference between males and females was observed in the reference group (Walk), but it was only significant in the Car group | ○ | ○ | ○ |
| When commuting from home to school neither main effect of commuting group, nor it's interactions with gender and commuting distance were significant | ○ | ✓ | ✓ |
| **Main scientific results** | ARX | SDV | SynDiffix |
| The main effect of commuting group on CRF and its interaction with distance were significant in the direction from school to home, but not in the opposite direction | ○ | ✗ | ○ |
| Predicted differences in CRF between commuting groups were moderate and generally higher in males than in females | ✓ | ✗ | ✓ |
| When comparing commuting group median distance from school to home, males driven by car had around 4 ml/ min/kg lower predicted CRF than those who walked or used wheels commuting (e.g., bicycle, skateboard) | ○ | ✗ | ○ |

As mentioned, besides the specific scientific statements, the paper suggests a policy intervention: that active transport should be promoted for children who use passive transport, but live within walking or biking distance to school. We regard this as the most important conclusion of the paper, and it is supported by ARX and SynDiffix, but would have been negated by SDV. The base paper starts with a number of simple statistical observations about the data (descriptive analytic results). Both ARX and SynDiffix support all six of these observations, while SDV negates four of them.

The main analytic thrust of the paper is the linear regression analysis of (CRF). Here, the performance of ARX and SynDiffix is less good. While none of the statement were negated, about half of the statements were not supported. While this would not have prevented the main policy conclusions from having been reached, the overall quality of the study would have suffered by using either anonymization technique.

### Usability of the anonymization tools and data

To enable data sharing in an open science environment, processes are needed that allow scientists to conveniently create and share safe data. As scientists already tend to be undermotivated to share data [2], it is hence beneficial if the anonymization step places as little additional burden on scientists as possible. Ideally, there should be little or no extra work required by data collectors or operators of repository systems to anonymize data and prepare them for sharing.

In terms of protected data generation, SynDiffix is the easiest to use of all methods studied here due to its simplicity and lack of configuration need. Using ARX, in contrast, requires expertise on the various anonymization methods that it supports, as well as substantial configuration efforts to generate a protected data table of sufficient quality. SDV is easier to use than ARX since it provides default configurations and code templates, but nevertheless requires that a decision about which SDV tool to use, and requires configuration of datatypes. However, in terms of data analysis, SynDiffix places an extra burden on the analyst, in that they need to understand that different tables need to be generated for different analytic tasks. Neither ARX nor SDV have this additional burden: the generated tables can be used as is.

In general, any anonymization technique requires that the analyst understands the extent to which data has been distorted, and requires that they compensate for that distortion. This always creates additional burden. Since the anonymization process in ARX is typically explainable and deterministic, and the present distortions are visible in univariate and bivariate statistics, they might be easier to understand. In contrast, typical synthetic data generation involves randomness and it is often not clear how the internal structure of the data might have changed, even if uni- and bivariate distributions appear to be similar.

## Discussion

In this paper, we compared the suitability of three tools for data anonymization (ARX, SDV and SynDiffix) for use in a public health study in Slovenia. Our study distinguishes itself from the large body of literature on this topic by studying in detail whether the individual analytical findings as well as the resulting conclusions would hold when using anonymized instead of the original data. Moreover, we explicitly considered the burden that would be imposed on researchers by applying one of the studied tools to follow open-science principles in our analysis.

Of the three tools, SDV’s data quality was poor and led to many incorrect scientific conclusions. The data quality of ARX and SynDiffix was good enough to provide definite value. The data generated by both tools supported the main conclusion of the base study (that children within walking or biking commuting distance should not commute with a car), and no incorrect conclusions were drawn. In addition, all of the descriptive analytics (counting and simple statistics) were supported by ARX and SynDiffix.

Regarding researcher burden for data generation, all three tools require some manual configuration. ARX typically requires configuration of data hierarchies per-column among other configuration choices, which requires expertise to find suitable settings. SDV requires selection of an algorithm, definition of data types, and potentially other configuration choices. If the dataset is time-series, SynDiffix only requires that the user configures the column that identifies individuals in the dataset. For the fitness dataset, ARX required around 200 lines of code (Java), SDV required 5 lines, and SynDiffix required 2 lines (both Python). Note that ARX is available as a GUI application (Windows, MacOS, or Linux), whereas SDV and SynDiffix require Python.

Regarding researcher burden for data analysis, the output of ARX and SDV can be used as is. SynDiffix generates multiple anonymized datasets, and the analyst must select the one with only the columns necessary for each given analytic task. This can only be done using a SynDiffix blob reader package that runs on Python. For the fitness study, a 22-line Python routine was needed to generate the two datasets used in a R script to generate Figure 3. In the analysis code, an extra line of code is required each time a dataframe is read.

A major limitation of the current study is that it does not truly replicate an open science scenario, whereby an analyst undertakes a scientific analysis purely from anonymized data. In the current study, the analyst had the advantage of hindsight, was already familiar with the raw data and its analysis, and simply replicated an analysis that had already taken place on the raw data. A more realistic study might be to give the anonymized data to a researcher unfamiliar with the data, have them undertake a scientific study from scratch, and then follow up the study with the raw data to determine not only whether the results are correct, but also whether they would have done the analysis differently given the raw data. This is a typical setting in secure processing environments, where researches might have only access to anonymized datasets on which they develop their analyses and run them separately on a protected environment.

Another limitation of this study is that it is based on only one dataset and analysis. The generalizability of findings might therefore be limited.

## Methods

### Choice of base study and dataset

The selection of the base study and corresponding dataset to use for this paper was limited to those in which the authors were involved. Besides the practical matter of having access to the original datasets, the authors of the original studies are in the best position to determine if the anonymized data is fit for purpose. We decided to select a base study where the dataset and the analysis are typical of those used by researchers participating in the SPOZNAJ^1^ open science project in Slovenia. Most such datasets are small, consisting of hundreds or a few thousand records. Using a small dataset also challenges the anonymization tools, since the perturbation introduced by anonymization has a proportionally stronger effect on small datasets.

In total, we considered eight studies [6, 10, 11, 12, 13, 14, 15, 16]. Of these, [10], [14], and [15] had too much data, and the analyses in [11] and [16] were less interesting than the others. Of the remaining studies, [6] was selected in part because it involves a sophisticated analysis technique (linear regression), and in part because it has a mix of significant and non-significant regression coefficients. An important and challenging test of anonymized data is how well it preserves significance.

### Choice of anonymized data methods

Anonymization mechanisms fall into two broad classes. One class aims to *replicate* or *modify* the original data as closely as possible so that statistical statements about the original data are accurate. Another class aims to *behave like* the original data in that they generate data suitable for predictive ML applications, and may even wish to modify the statistics of the data, by for instance creating additional data, possibly with bias added or removed. This latter class is generally referred to as “synthetic data”, although there is no strict definition of this term[17].

There is a long history of open source tools in the replicate class, including sdcMicro [18] which implements techniques classically used by statistics agencies (swapping, outlier removal, sampling), ARX [19] which implements *k*-anonymity [20] and related techniques among others, Synthpop [21] which implements the decision tree approach CART [22] among others, and most recently SynDiffix [8] which is a multi-table tree-based approach.

Of these, we selected ARX which has demonstrated success particularly in medical domains [19, 23], and SynDiffix which claims high accuracy and ease of use [24]. The anonymized data generated by both of these tools is row-level data (also known as microdata) that syntactically is equivalent to the original data. In this specific narrow sense, ARX and SynDiffix can be thought of as synthetic data.

We wished also to select at least one tool from the behave-like class. These techniques have received considerable attention in recent years since the publication of the CTGAN and TVAE techniques [25], and a number of open source and commercial tools are available. We tested both the open source tool Synthetic Data Vault (SDV)[9] and the commercial product Mostly AI. The two demonstrated similar results for this paper’s dataset, so we selected SDV since open source is better for open science.

Note that the anonymized datasets generated for this paper for ARX and SynDiffix were built by the respective authors of those tools. The SDV dataset was generated by Francis.

The procedures used to generate the three anonymized datasets from ARX, SDV, and SynDiffix, are described in the following sections.

### ARX

#### Overview

The ARX software is intended to anonymize sensitive personal data and supports a wide variety of privacy models and data transformation methods. It can be used either with a graphical user interface as a standalone software, or by using the Java-based ARX library to perform the data anonymization via code [23]. ARX allows for fine-grained configuration to implement tailored anonymization procedures and offers a wide variety of options to protect the data while providing high performing optimization algorithms to retain the data utility [19].

#### Anonymized data generation

To perform the data anonymization, ARX requires dataset-specific configuration. This includes the privacy models to be used and their thresholds as well as domain-generalization hierarchies for the variables that can be used by the software to aggregate data tailored to the scientific research question or as a distance-measure during clustering. Finally the specific transformations to be performed, e.g., suppression, full-domain or local generalization, aggregation, as well as the algorithm used to perform the optimization process need to be specified. Parameters often need to be fine-tuned over several iterations.

For the given dataset, the process was quite straight forward. We chose *k*-Anonymity as a strict privacy model that protects all records and applies to all variables. We chose the threshold *k*=2, because it is the weakest possible parameterization and weaker parameters tend to provide higher utility for small datasets. ARX was configured to perform a clustering process where domain-generalization hierarchies are used to determine distances between values. For categorical variables, a simple domain-generalization hierarchy was designed, grouping “walk” and “wheels” together, because they both indicate movement through physical activity. For continuous variables, the associated domain-generalization hierarchies represented increasingly large intervals. The hierarchy for the “gender” variable simply included a common root node “*” for both genders. In each cluster, categorical values were replaced by the mode of all values in the cluster, while drawing from the distribution within input data if there was no mode, and continuous variables were replaced with the arithmetic mean of all values in the cluster.

We applied the transformations using ARX’s local optimization strategy [19] using the Java library provided by ARX version 3.9.2. The core Java code (i.e. excluding I/O, imports, etc.) for executing the anonymization is around 200 lines. Executing the process took about 2.3 minutes on a laptop with an i5-1135G7 processor running at 2.4GHz and sufficient memory (8GB).

#### Anonymized data usage

The data anonymization performed by ARX retained the data structure of the original data, therefore no post-processing was needed before use.

### Synthetic Data Vault (SDV)

#### Overview

The Synthetic Data Vault (SDV) is an open source project implementing several synthetic data toolscitepatki2016synthetic. SDV offers different tools depending on whether the original table is a single table, multiple tables (relational), or time-series.

The single-table case offers several tools, such as Gaussian-Copula, CTGAN, and TVAE. The latter two are promoted by SDV as being suitable for data with a mix of categorical and continuous columns, and the data quality of CTGAN and TVAE is similar [25].

We used SDV’s CTGAN tool (Conditional Tabular Generative Adversarial Network) to generate an anonymized dataset. As the name implies, CTGAN uses a Generative Adversarial Network (GAN) approach [25]. It runs two neural networks, a *generator* and a *discriminator*. The generator tries to create anonymized datasets that the discriminator cannot distinguish from the original data, while avoiding overfitting.

There are a number of parameters that can be used to fine-tune CTGAN. Most importantly, the metadata description must be correct, especially the labeling of continuous and categorical columns. There are a number of other parameters related to the GAN itself (e.g., enforced minimums and maximums, enforced rounding, number of epochs) that can improve data quality somewhat.

#### Anonymized data generation

SDV has an option to auto-generate the metadata. We used this option and checked to ensure that the metadata was correct. Since all of the categorical columns in the original dataset are strings, the auto-generated metadata was correct. Since simplicity of operation is an important requirement in an open science environment, we chose to use the default parameter settings. Note that the commercial synthetic data product Mostly AI, which has sophisticated algorithms to automate the selection of models and parameters and in general outperforms SDV’s CTGAN [24], did not in this case perform better than CTGAN with the default settings. We therefore believe that we could not have improved substantially by tweaking the parameters.

~~~
from sdv.metadata import SingleTableMetadata
from sdv.single_table import CTGANSynthesizer
metadata = SingleTableMetadata()
metadata.detect_from_dataframe(df_orig)
synthesizer = CTGANSynthesizer(metadata)
synthesizer.fit(df_orig)
df_syn = synthesizer.sample(num_rows=len(df_orig))
~~~

The core Python code is five lines. We used version 1.14.0 of SDV with the default settings. It took 32 seconds to generate the anonymized data on a laptop with an i7-7820HQ processor running at 2.9GHz and sufficient memory (32GB).

#### Anonymized data usage

The data anonymization performed by SDV retains the data structure of the original data, therefore no post-processing is needed before use.

### SynDiffix

#### Overview

SynDiffix takes a *multi-table* approach to synthesizing data [8]. A key characteristic of all data anonymization methods is that accuracy degrades as the number of columns increases. Therefore it is better to synthesize only those columns needed for a given analytic task. Given that there can be thousands of different combinations of columns that analysts may be interested in, a multi-table approach can lead to the generation of thousands of distinct tables. Unlike other anonymized data methods, SynDiffix is designed to maintain anonymity no matter how many anonymized tables are generated.

When SynDiffix synthesizes a table with more than around 5 or 6 columns, SynDiffix partitions the table into a set of sub-tables each with fewer columns, synthesizes each sub-table individually, and then joins the sub-tables back together. Columns within sub-tables are more strongly correlated than columns across sub-tables. Each sub-table has at least one column in common with another sub-table, and these common columns are used for joining.

SynDiffix is implemented in Python, and has two modes of operation, *full-table* mode and *sub-table blob* mode. In full-table mode, the user (data controller or analyst) requests a single table, and SynDiffix returns that table after doing any required partitioning and joining. Full-table mode requires that the original data is available to SynDiffix. Full-table mode uses the `Synthesizer` class of SynDiffix.

In sub-table blob mode, a single API call is made by the data controller to create the complete set of sub-tables required to generate any table. This set of sub-tables is zipped into a single file which is refered to as a *SynDiffix blob*. The blob may safely be released to the public. Creating the blob uses the `SyndiffixBlobBuilder` class, and requires access to the original data. Subsequently, an analyst with access to a blob uses the `SyndiffixBlobReader` class to join and retrieve tables from the blob. The analyst requests a table with the given columns, the blob reader selects the appropriate sub-tables from the blob, joins them together, and returns the table. The original data is not required for this operation.

In either mode, no table-specific configuration is required if the table is not longitudinal or time-series. If the table is, then the column that identifies each individual in the dataset must be specified. No other configuration is necessary.

#### Anonymized data generation

To generate the blob, the user must write a python script to read in the original data file as a dataframe (`df_original` below), and generate and save the blob. Generating the blob required two lines of Python:

~~~
sbb = SyndiffixBlobBuilder(blob_name, blob_path)
sbb.write(df_original)
~~~

This creates a blob from the full original dataframe `df_original` and places it in the directory at `blob_path` with the name `blob_name`.

The blob file created from the original data with eight columns and 713 records contains 160 tables, took 81 seconds to build on the same laptop as with ARX, and is 1.4MB in size (zipped).

#### Anonymized data usage

While creating the blob is simple and automatic and requires no expertise from the controller, using the blob for analytics is unfortunately more complex. For each analytic task, the analyst must request from the blob a table with only the required columns. In addition, if the analytic task is to build a predictive model for a given target column, then the target column should be specified in the blob request. The analyst must be aware of these requirement, and must modify their analysis code to satisfy the requirements. The quality of the data is substantially worse if the analyst instead uses a single full table for all of their analysis.

Recreating the three tables and one plot from the original paper required seven different tables from the blob; 2 with 1 column, 3 with 2 columns, and 2 with 6 columns and a target column (VO_2_max). In the scripts used for generating the tables and plot, an additional API call to `SyndiffixBlobReader` was required prior to each analytic task:

~~~
sbr = SyndiffixBlobReader(blob_name, blob_path)
df1 = sbr.read([col1, col2]) analytic_task1(df1)
df2 = sbr.read([col3, col4, col5, col6], target_column=col5)
analytic_task2(df2)
…
~~~

## Author contributions statement

P.F. generated the SynDiffix and SDV datasets, and generated the paper’s tables and figures. K.O. and F.P. generated the ARX dataset. B.L. and G.J. interpreted the validity of the anonymized datasets relative to the original data. P.F. drafted the initial version of the manuscript. All authors reviewed and edited the manuscript.

## Data Availability

The repository at https://github.com/yoid2000/commute-health-study contains all of the anonymized data. Note that the original data contains personal data and is therefore not publicly available.

## Acknowledgements

F.P. and K.O. acknowledge funding from the German Ministry of Health (project KI-FDZ, grant agreement number 2521DAT01C) and from the German Research Foundation (project NFDI4Health, project number 442326535). This research has been co-financed by the Horizon Europe programme of the European Union within the framework of the SmartCHANGE project (nº 101080965), and the Slovenian Research and Innovation Agency (Bio-psycho-social research program, nº P5-0142).

## Code Availability

### Accession codes

The repository at https://github.com/yoid2000/commute-health-study contains:

- The Python code used to generate the SDV and SynDiffix anonymized data.
- The `.jar` executable to generate the ARX anonymized data.
- All of the anonymized data.
- The R script and Python code used to generate all of the tables and figures in this paper.

The repository at https://github.com/BIH-MI/commute-health-anonymization contains the source code (Java) to generate the ARX anonymized data.

## Footnotes

1 https://projekt-spoznaj.si/

